# Screening and vaccination against COVID-19 to minimize school closure

**DOI:** 10.1101/2021.08.15.21261243

**Authors:** Elisabetta Colosi, Giulia Bassignana, Diego Andrés Contreras, Canelle Poirier, Pierre-Yves Boëlle, Simon Cauchemez, Yazdan Yazdanpanah, Bruno Lina, Arnaud Fontanet, Alain Barrat, Vittoria Colizza

## Abstract

Schools were closed extensively in 2020-2021 to counter COVID-19 spread, impacting students’ education and well-being. With highly contagious variants expanding in Europe, safe options to maintain schools open are urgently needed. We developed an agent-based model of SARS-CoV-2 transmission in school. We used empirical contact data in a primary and a secondary school, and data from pilot screenings in 683 schools during the 2021 spring Alpha wave in France. We fitted the model to observed school prevalence to estimate the school-specific reproductive number and performed a cost-benefit analysis examining different intervention protocols. We estimated R^Alpha^=1.40 (95%CI 1.35-1.45) in the primary and R^Alpha^=1.46 (1.41-1.51) in the secondary school during the wave, higher than R_t_ estimated from community surveillance. Considering the Delta variant and vaccination coverage in Europe, we estimated R^Delta^=1.66 (1.60-1.71) and R^Delta^=1.10 (1.06-1.14) in the two settings, respectively. Under these conditions, weekly screening with 75% adherence would reduce cases by 34% (95%CI 32-36%) in the primary and 36% (35-39%) in the secondary school compared to symptom-based testing. Insufficient adherence was recorded in pilot screening (median ≤53%). Regular screening would also reduce student-days lost up to 80% compared to reactive closure. Moderate vaccination coverage in students would still benefit from regular screening for additional control (23% case reduction with 50% vaccinated children). COVID-19 pandemic will likely continue to pose a risk for school opening. Extending vaccination coverage in students, complemented by regular testing largely incentivizing adherence, are essential steps to keep schools open, especially under the threat of more contagious variants.

## INTRODUCTION

School closure has been extensively used worldwide against the COVID-19 pandemic. The first wave witnessed many countries go into strict lockdowns closing schools for long periods of time [1], and their reopening has been continuously challenged by successive waves and the need for social distancing restrictions. In Europe, depending on the country, students lost from 10 to almost 40 weeks of school from March 2020 to March 2021 due to partial or total school closure (Figure 1a). Strategies were affected by the limited understanding of viral circulation in children and their contribution to transmission [2].

**Figure 1.**
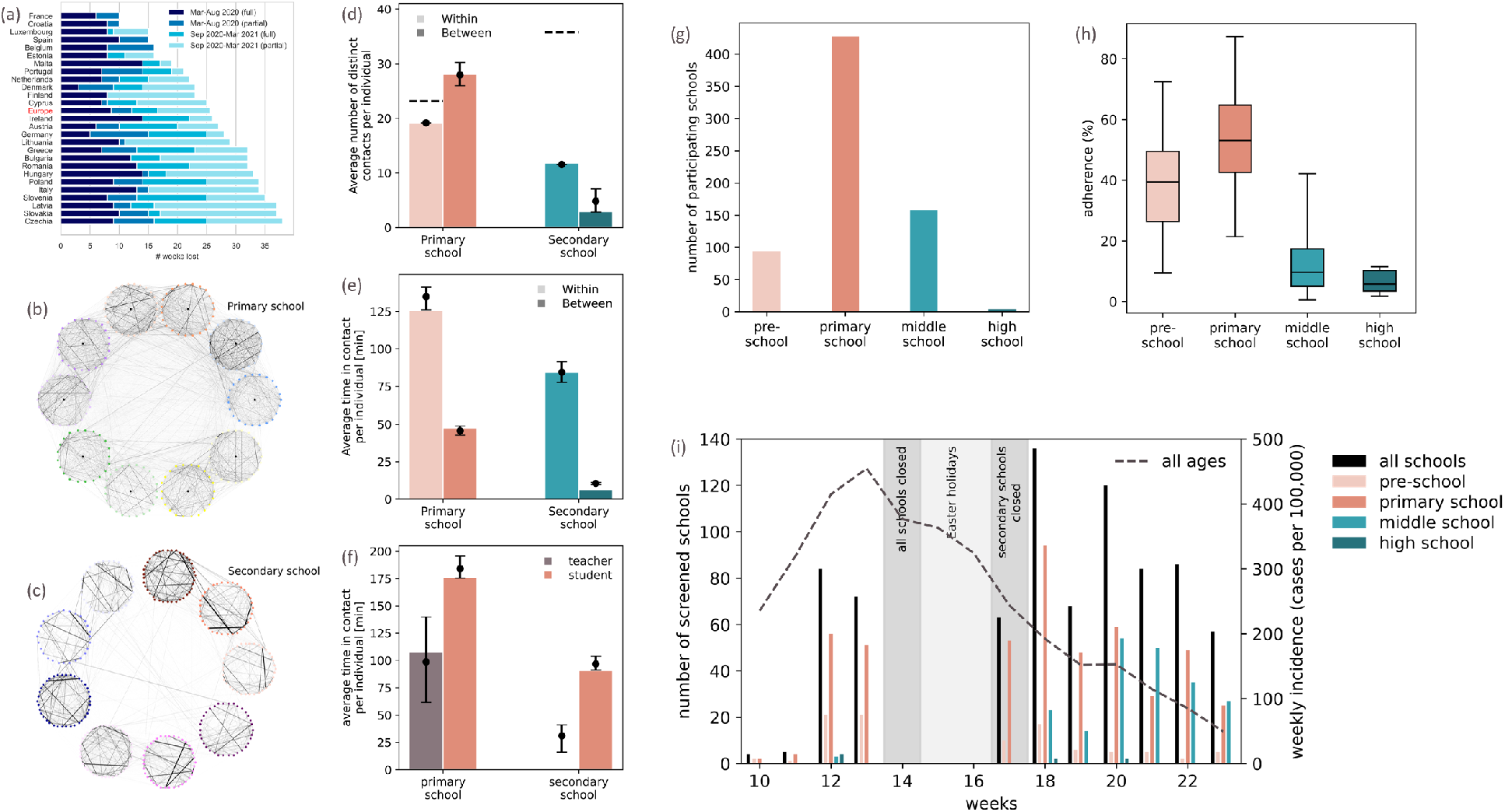
School closure in Europe, empirical contact networks in a primary school and a secondary school, and field screening data in schools in France. (a) Average number of in-presence school weeks lost by students in Europe because of school closures due to the pandemic. Source: Unesco [1]. (b), (c) Visualization of the empirical temporal contact data aggregated over two days, for the primary (panel b) and the secondary (panel c) school. Nodes represent teachers and students, each circle represents a class (each of a different color), and links represent contacts, with the thickness coding the contact duration. In the secondary school classes are divided in three groups based on the specialization (mathematics and physics; physics, chemistry, engineering studies; biology). (d) Daily average number of distinct contacts per individual within the same class or in different classes, in the primary and secondary school. Horizontal dashed lines represent the average class size. (e) Daily average time that an individual spends in interaction within the same class or in different classes, in the primary and secondary school. (f) Daily average time that an individual spends in interaction for teachers and students in the primary school. In panels d-e-f, histogram bars refer to the empirical networks. Points and error bars (95% bootstrap confidence intervals) refer to the synthetic networks. In panels d-e, the increase of average number of contacts and duration in the synthetic secondary school networks compared to their empirical counterparts is due to the *ad hoc* addition of contacts between school years. In panel f, no empirical data is shown for teachers, as they did not participate to the data collection, and their contact behavior was inferred from another dataset. (g) Number of schools participating to the pilot screenings initiative during the spring 2021 wave in the Ain, Loire, and Rhône departments according to the school level. (h) Observed adherence to screening recorded in the different school levels participating to the pilot screenings. Box plots represent the median (line in the middle of the box), interquartile range (box limits) and 2.5th and 97.5th percentiles (whiskers). (i) Weekly incidence over time (cases per 100,000, right y axis) from community surveillance in the 3 departments under study (Ain, Loire, Rhône; SI, Section 2) and number of schools participating to the pilot screenings (left y axis) over time by school type during the 2021 spring wave in France. The vertical shaded areas indicate the school closures in the period under study.

Outbreaks in schools are difficult to document, as infections in children are mostly asymptomatic or present mild non-specific symptoms [3]. Despite the lower susceptibility to infections in children compared to adults [4], viral circulation can occur in school settings, especially in secondary schools [2]. Accumulating evidence is consistent with increased transmission in the community if schools are in session [2], [5], and model-based findings suggest that school closure may be used as an additional brake against the COVID-19 pandemic if other social distancing options are exhausted or undesired [6], [7].

Keeping schools safely open remains a primary objective that goes beyond educational needs, and affects the social and mental development of children [8], as well as the reduction of inequalities. Several countries implemented safety protocols at school, including the use of masks, hand hygiene, staggered arrival and breaks. Regular testing [9]–[12] was introduced in a few countries as an additional control measure. Vaccination was extended to the 12+ population and recently approved for children in Europe, yet it is unlikely that primary schools will be largely vaccinated during the 2021-2022 winter. The rapid surge of cases reported in Europe at the time of writing due to the Delta variant [13] threatens classroom safety. The risk is further exacerbated by the emergence of novel variants [14]. Assessing vaccination and protocols in schools is therefore key to maintaining schools open. Here, through an agent-based transmission model parameterized on empirical contacts at schools and fitted to field screening data in schools, we estimate the school-specific effective reproductive number. We then evaluate intervention protocols combining closures and screening, under varying immunity profiles of the school population, and accounting for age-specific differences in susceptibility to infection, contagiousness, contact patterns, and vaccine effectiveness.

## METHODS

### Empirical patterns of contacts

We used empirical data describing time-resolved face-to-face proximity contacts between individuals in two educational settings, collected in France using wearable RFID sensors in a pre-pandemic period. The *Primary school* dataset describes the contacts among 232 students (6-11 years old) and 10 teachers in a primary school in Lyon, composed of 5 grades, each of two classes [15]. The *Secondary school* dataset describes the contacts between 325 students (17-18 years old) of 9 classes in a secondary school in Marseille [16]. Classes belong to the second year of “classes préparatoires”, specific to the French schooling system for preparation to University entry, and are divided in three groups, based on the specialization.

We built temporal contact networks, composed of nodes representing individuals (classified by class and student/teacher), and links representing empirically measured proximity contacts occurring at a given time (Figure 1b,c). As each dataset covers only a few days, we developed an approach to temporally extend the datasets by generating synthetic networks of contacts that reproduce the main features observed empirically (class structure, within- vs. between-classes links, contact duration heterogeneity, and similarity across days). The secondary school synthetic network was further extended to generate a synthetic first year (to consider the full curriculum of the “classes préparatoires”) including teachers whose contacts were inferred from an additional dataset for the same school. The resulting network for the secondary school was composed of 650 students and 18 teachers (Supplementary Information (SI), Section 5).

### Field screening data in schools during the spring 2021 wave in France

In response to a rising third wave in France in the spring 2021 due to the Alpha variant, local authorities in the Auvergne-Rhône-Alpes region proposed pilot screenings at schools on a voluntary basis to detect cases. We used data on adherence to screening and test results collected in 683 schools between March 8 and June 7, 2021 (weeks 10-23), in the Ain, Loire and Rhône departments of the region. Screening was interrupted in April due to reactive school closure (week 14) and Easter holidays (weeks 15-16) while the country underwent the third national lockdown; it was resumed in week 17 at school reopening (week 18 for secondary schools; Figure 1i). Screenings involved 94 pre-schools, 427 primary schools, 158 middle schools, and 4 high schools, for a total of 209,564 students and 18,019 personnel tested. PCR tests from saliva samples were proposed in pre-schools and primary schools, and anterior nasal LFD (lateral flow device) tests in middle and high schools. More details are provided in the SI (Section 2).

### Ethics statement

Contact studies were approved by the Commission Nationale de l’Informatique et des Libertés (CNIL, the French national body responsible for ethics and privacy) and school authorities. Informed consent was obtained from participants or their parents if minors. No personal information of participants was associated with the RFID identifier. Testing at school was part of surveillance activities approved by school authorities and proposed with parental consent. Screening data were provided in aggregated and anonymized form.

### Transmission model in primary school and secondary school

We developed a stochastic agent-based model of SARS-CoV-2 transmission on the network of contacts. Infection progression includes prodromic transmission, followed by clinical or subclinical disease stages, informed from empirical distributions. Transmission occurs with a given transmissibility *β* per contact per unit time between an infectious individual and a susceptible one. *β* was inferred by fitting the model to data from screening results during the 2021 spring wave. Individuals in the asymptomatic compartments are considered to be less infectious and to remain undocumented unless tested [17]; a sensitivity analysis was performed on the reduced transmissibility.

The model is parameterized with age-specific estimates of susceptibility, transmissibility, probability of developing symptoms, and probability to detect a case based on symptoms (SI, Section 1). A systematic review indicates that minors have lower susceptibility to SARS-CoV-2 compared to adults [4], but building evidence suggests that high school students may be as susceptible as adults [18]. Here we considered a relative susceptibility of 50% in children and 75% in adolescents compared to adults, and tested 100% susceptibility in adolescents for sensitivity. The probability to recognize a suspect COVID-19 infection from symptoms was set to 30% for children and 50% for adolescents and adults, based on studies indicating that about two thirds of symptomatic children [3] and half of symptomatic adults [19] have unrecognized symptoms before diagnosis. These values were varied for sensitivity. We considered a lower transmissibility in children, as evidence suggests that transmission in children may be less efficient [20], and we tested different values for sensitivity.

The model is further stratified to account for vaccination status and to include vaccine effectiveness against infection, transmission, and clinical symptoms given infection, accounting for waning protection [21] (SI, subsection 1.4). Higher and lower vaccine effectiveness were also tested for sensitivity. Full details on the transmission model are reported in the SI (Section 1).

### Closure and screening protocols

*Symptom-based testing and case isolation (ST)* is considered as the basic strategy, present in all protocols, and against which interventions are evaluated. It considers that clinical infections are detected with the estimated probability and tested, and confirmed cases are isolated for 7 days. We considered the following intervention protocols:

- *Reactive quarantine of the class (ST+Qc):* once a case is identified through ST, their class is put in quarantine for 7 days. If quarantined individuals develop symptoms, they remain in isolation for an additional period of 7 days, before returning to school. This protocol was largely adopted in France throughout the pandemic.
- *Reactive quarantine of the class level or specialization (ST+Ql):* as the previous protocol, but quarantine is applied to the classes of the same level (2 classes in the primary school) or specialization (3 in the secondary school) of the detected case. This option is considered as empirical data show a larger mixing between students of the same level or specialization compared to the others.
- *Reactive screening of the class (+1d from detection) followed by a control screening (+nd) with α adherence (ST+rT+cnTα%)*: once a case is identified through ST, their class is reactively screened at +1 day, and again at +n days (n=4, or 7) for control of possible infections that went previously undetected. Only a percentage *α* of the non-vaccinated school population adheres to the screening.
- *Regular testing with α adherence (ST+RTα%):* in addition to ST, regular testing is performed at a certain frequency (once every two weeks, once or twice per week). Adherence *α* was informed from field data, and further explored in a range between 10% and 100%.
- *Regular testing with α adherence, and reactive quarantine of the class (ST+RTα%+Qc):* in addition to the protocol above, the reactive closure of the class is triggered at every detected case.

Following protocols adopted in France, we considered PCR tests on saliva samples in the primary and anterior nasal LFD tests in the secondary school, with time-varying test sensitivity specific to each test, and results available after 24h and after 15’, respectively (SI, subsection 1.4). Teachers are required to show proof of a negative PCR test when returning to school after infection.

### Inference framework

We used data on test results collected in the pilot screenings during the 2021 spring wave in the Ain, Loire and Rhône departments to estimate the transmissibility *β*^*Alpha*^ per contact per unit time of the Alpha variant and the corresponding school-specific effective reproductive number R in that period. The model is fitted to the observed prevalence of cases in students in the tested schools through a maximum likelihood approach. We used data from screenings performed during the rise of the spring wave (March 8 to April 2, 2021), involving at least 5 schools and 500 screened students per week per department per school type (primary or secondary), and with reported adherence ≥50% (reference inclusion criteria). For sensitivity, we relaxed the constraint on adherence (sensitivity inclusion criteria). Simulations for the fit covered the period from week 8 (starting February 22, 2021, at school reopening after winter holidays) to week 13 (ending April 4) before the reactive school closure, and they were initialized with age-specific seroprevalence estimates [22]. Weekly introductions at school were modeled stochastically, inferred from age-specific community surveillance data, and adjusted to account for detection rate and within-school transmission [23]. We computed R in each school as the ratio of the number of individuals infected at the 2nd generation to the number infected at the 1st generation for each initial seed over 5,000 simulated outbreaks. The estimated R refers to the ST+Qc protocol with mask mandate applied in that period. Full details on the procedure are reported in the SI (Section 3).

### Analysis of school protocols in a Delta winter scenario in Europe

To evaluate the efficacy of intervention protocols, we considered a 2021-2022 winter scenario due to the Delta variant initialized with 25% natural immunity in the population, 60% of teachers vaccinated, and 40% of adolescents vaccinated, corresponding to the median vaccination coverage registered in countries in Europe by mid-September 2021 (SI, Section 4). The transmissibility *β*^*Delta*^ per contact per unit time for Delta was estimated from the maximum likelihood estimate *β*_*MLE*_ = *β*^*Alpha*^, accounting for the transmissibility advantage of the Delta variant [24]. The corresponding school-specific R was estimated from simulated outbreaks under the above immunity conditions, and considering the ST+Qc protocol with mask mandate. We additionally explored a range of R values to account for the uncertainty in the estimate of Delta transmissibility [24], seasonal effects [25], and variations in *β*_*MLE*_ due to the inclusion criteria considered in the inference. We considered low, moderate, sustained, and high weekly introductions modeled stochastically and corresponding to community surveillance incidence in primary school students ranging in time from 25 to >600 cases per 100,000 (low introductions), from 50 to 900 (moderate), from 100 to 1,300 (sustained), and from 200 to 1,800 cases per 100,000 (high); values for the secondary school are reported in the SI (subsection 4.2).

To assess the efficacy of screening protocols under different immunity conditions, we explored a full range of vaccination coverage in children, adolescents, and teachers.

### Simulation details and analysis

Estimates for *β* and R were obtained from 5,000 simulated stochastic outbreaks for each parameter set. Estimates for R were compared to age-specific R_t_ estimated from community surveillance data with a one-sample t-test. We fitted the predicted offspring distribution to a negative-binomial to estimate the overdispersion parameter k [26]. In the protocols’ analysis, we performed 1,000 stochastic runs for the primary and 2,000 for the secondary school for each parameter set, over the course of a trimester (90 days). We computed medians and 95% bootstrap confidence intervals from simulation outputs to compare protocols with a Mood’s median test. Interquartile ranges (IQR) were used to describe observed adherence.

## RESULTS

Contact networks measured through wearable sensors displayed a strong community structure around the classes, common to both the primary and secondary schools (Figure 1b,c). The patterns of interaction, however, varied substantially between the two settings. On average, children had a larger number of distinct contacts during a day, interacting with almost their entire class (83% of the class), compared to adolescents (33% of the class, Student test p<10^−15^; Figure 1d). Approximately 50% more links occurred between classes than within classes in the primary school (19 vs. 28 links, p<10^−15^), contrary to what observed for adolescents (12 vs. 3 links, 75% fewer links, p<10^−15^). But accounting for duration, students in both settings spent on average more time interacting within the class than outside the class (p<10^−15^), and established longer contacts (+64%, p=0.009) compared to teachers (Figure 1e,f).

Using the empirical contact patterns, we inferred the school-specific transmissibility from screening data in primary schools satisfying the inclusion criteria: 71 primary schools and 12,146 tested students with the reference inclusion criteria; 103 primary schools and 15,916 tested students with the sensitivity inclusion criteria. Secondary schools were excluded because of limited participation. We estimated a school-specific R^Alpha^ during the Alpha 2021 spring wave in France between 1.40 (1.35 −1.45) and 1.44 (1.40-1.48) in the primary school, and 1.46 (1.41 - 1.51) and 1.50 (1.46-1.54) in the secondary school (for the reference and sensitivity inclusion criteria, respectively), with the reactive class closure protocol and mask mandate in place (Figure 2a). Estimates were higher compared to the time-varying reproductive number R_t_ obtained from age-specific community surveillance in the same period (one-sample t-test p<10^−7^ in the primary, p=3·10^−5^ in the secondary school; Figure 2c,d). We quantified a large individual-level variation in SARS-CoV-2 transmission in both schools, corresponding to an overdispersion parameter k estimated to be 0.56 (95% CI 0.49-0.63) in the primary and 0.52 (95% CI 0.46-0.58) in the secondary school (Figure 2b). Accounting for the transmissibility advantage of the Delta variant and vaccination coverage in Europe, we estimated a school-specific R^Delta^ between 1.66 (1.60 −1.71) and 1.70 (1.66-1.75) in the primary school, and 1.10 (1.06 −1.14) and 1.13 (1.10-1.16) in the secondary school (for both inclusion criteria). In the protocols’ analysis, we considered the R^Delta^ estimate obtained with the reference inclusion criteria, and explored the ranges 1.46-2.00 and 0.97-1.34 in the primary and secondary schools, respectively, estimated accounting for the uncertainty associated to Delta transmissibility, seasonal effects, and sensitivity inclusion criteria.

**Figure 2.**
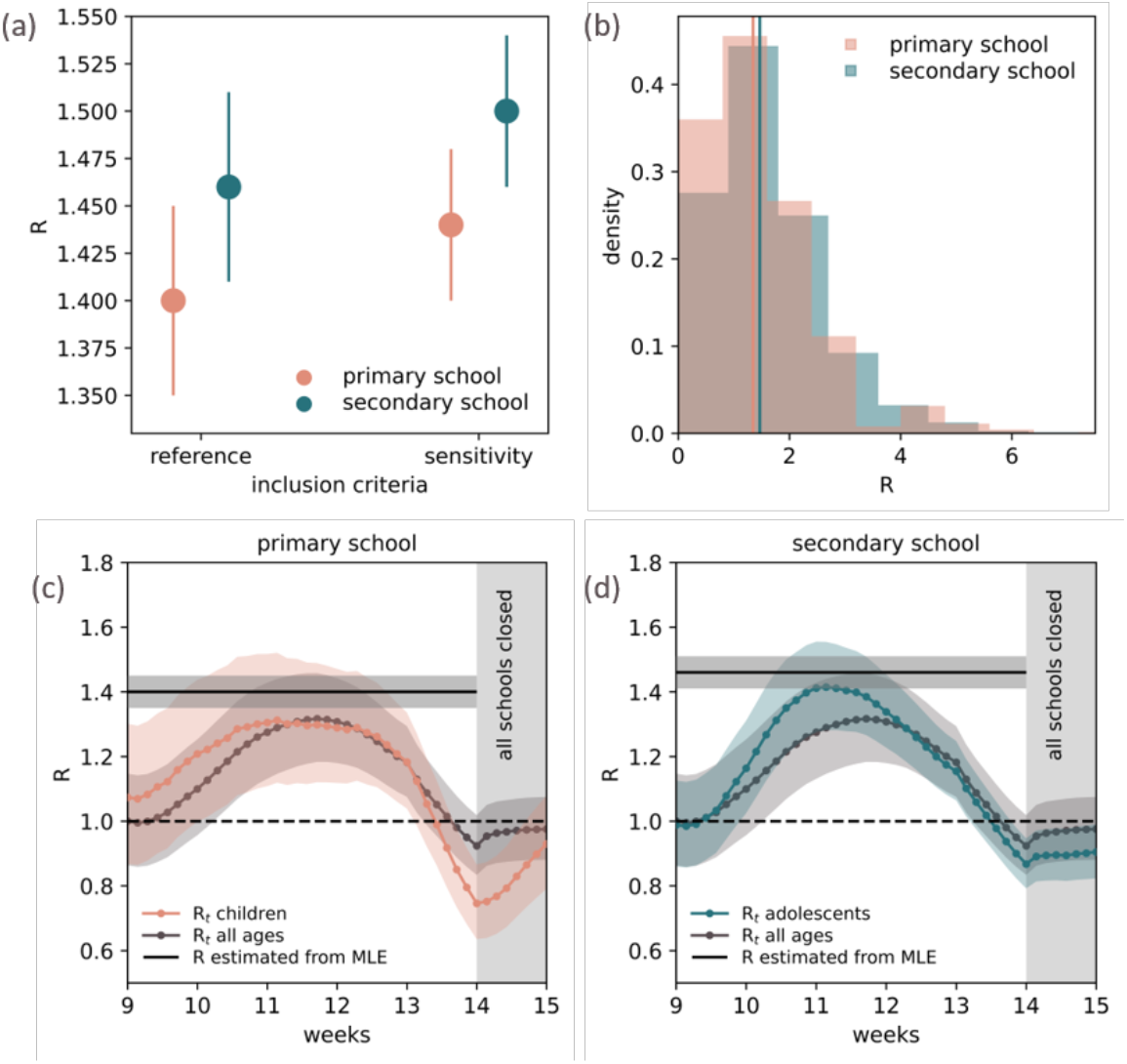
Estimates of the effective reproductive number in the school setting during the 2021 spring wave in France due to the Alpha variant. (a) Estimates of the effective reproductive number in the primary school and secondary school obtained with the reference inclusion criteria and the sensitivity inclusion criteria. Estimates refer to the Alpha variant during the 2021 spring wave in France, when reactive closure of the class and mask mandates were in place. Errors indicate the 95% confidence intervals. The estimates are obtained from the maximum likelihood estimates (MLEs) of the transmissibility *β* = *β*_*MLE*_ per contact per unit time by fitting the agent-based transmission model to observed prevalence in schools in the pilot screenings. (b) Predicted offspring distribution in the primary and secondary school. Vertical lines indicate the effective reproductive number (i.e. the average of the distribution) in each school obtained with the reference inclusion criteria. (c) Comparison between the model estimate of the effective reproductive number (R estimated from the MLE of the transmissibility *β* in the primary school, horizontal line; the shaded area corresponds to its 95% confidence interval) and the time-varying reproductive number *R*_*t*_ estimated from community surveillance incidence in children (orange) in the three departments under study during the rise of the 2021 spring wave (SI, subsection 3.5); *R*_*t*_ for all ages (grey) is shown for reference. The shaded area around the *R*_*t*_ curve indicates the 95% credible intervals. (d) As in panel c, for the secondary school.

Under the estimated Delta transmissibility and sustained introductions, regular testing constitutes an efficient protocol for preventing infections in a partially immunized school population (Figure 3a,b). If adherence is large enough, regular testing can substantially outperform protocols based on simply identifying cases given recognizable symptoms and additionally closing or screening the class of the detected case (even with a follow-up control screening). However, screenings at schools during the 2021 spring wave in France were met with low or moderate participation rates. Adherence was higher in lower school levels (39% (IQR 26-49%) in pre-school, 53% (43-65%) in primary school) compared to secondary schools (10% (5-17%) in middle school, 6% (3-10%) in high school; Mood’s median test p<10^−15^; Figure 1h). We found that with 50% adherence, i.e. approximately the value recorded in the French primary schools, weekly screening would reduce the number of cases by 21% (95%CI 19-23%) in the primary and by 26% (25-28%) in the secondary school compared to symptom-based testing alone. Case reduction would rise to 34% (32-36%) and 36% (35-39%) in the two schools, respectively, with 75% adherence.

**Figure 3.**
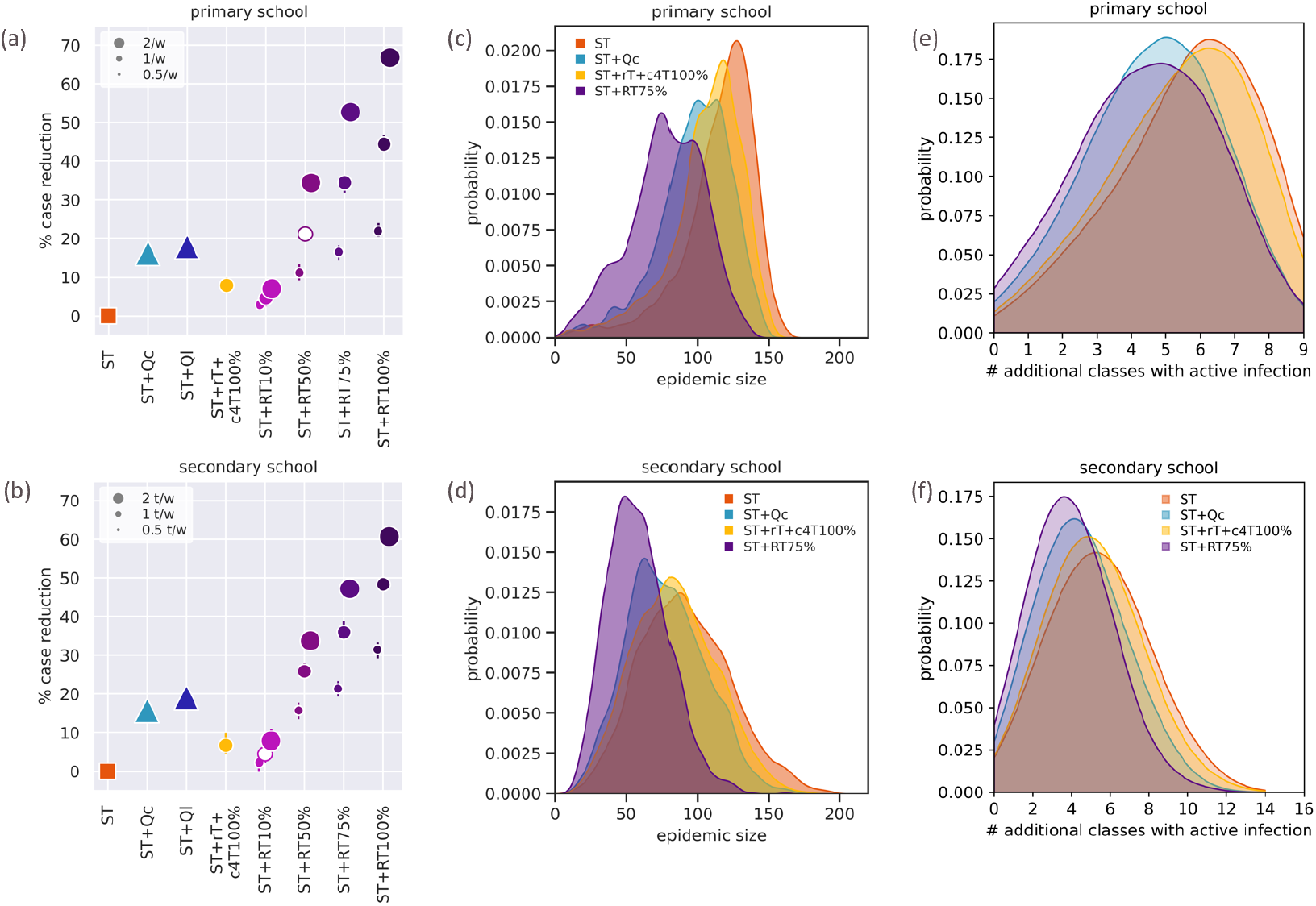
Efficiency of regular testing in educational environments. (a) Predicted percentage of reduction in the number of cases achieved by each intervention protocol with respect to the basic strategy of the symptom-based testing and case isolation (ST) in the primary school. The reduction is computed on the final size over the timeframe of 90 days. Intervention protocols are: symptom-based testing with reactive quarantine of the class (ST+Qc); symptom-based testing with reactive quarantine of the class level (ST+Ql); symptom-based testing and reactive screening of the class (+1d from detection), followed by a control screening (+4d) with adherence α=100% (ST+rT+c4T100%); symptom-based testing coupled with regular testing (ST+RTα%) with adherence α=10%, 50%, 75%, 100%. For regular testing, different frequencies are shown: one test every two weeks, a weekly test, two tests per week. Error bars correspond to 95% bootstrap confidence intervals; in some cases they are smaller than the symbol size. The empty marker corresponds to the adherence estimated from empirical data recorded in primary schools. Additional results for the reactive screening are reported in the SI, subsection 6.7. (b) As in panel a for the secondary school. (c) Probability distribution of the simulated final epidemic size in the primary school for selected protocols over the timeframe of 90 days. Four selected protocols are shown. Regular testing is done with weekly frequency. (d) As in panel c, for the secondary school. (e) Probability distribution of the additional number of classes in the primary school with at least one active infection when a case is confirmed. Four selected protocols are shown. Regular testing is done with weekly frequency. (f) As in panel e, for the secondary school. In all panels, simulations are parameterized with sustained introductions and the estimated effective reproductive number for the Delta variant, R=1.66 in the primary and R=1.10 in the secondary school, accounting for differences in vaccination coverage. R refers to the ST+Qc protocol with mask mandate, corresponding to the conditions applied in France when data used for the inference were collected.

Alternatively, similar reductions would be achieved with 50% adherence and twice-weekly testing. This shows how infection prevention improves with both adherence and frequency of tests, and higher frequency is needed to compensate for lower adherence. However, if adherence to regular testing is too low (10%), as recorded in the French secondary schools, weekly testing would have little impact (<10% case reduction), similarly to reactive screening and lower than reactive closure. While trends are similar across settings, partial vaccination coverage in adolescents leads to smaller epidemic sizes in the secondary school compared to the primary (relative to the school size; Figure 3c,d and SI, subsection 6.5).

Next to reducing the number of infections, regular testing is predicted to strongly limit the number of days of absence of students. The quarantine of the class implies 17.7 (95% CI 17.4-17.9) and 33 (95% CI 32-34) times more student-days lost in the primary and secondary schools, respectively, compared to symptom-based testing alone (Figure 4a). Days lost inevitably increase when reactive closure is extended to classes of the same level or specialization. Not being sufficiently targeted, reactive closure quarantines individuals while their risk of infection may be low, and the virus may have spread to other classes (Figure 3e,f). Reducing mixing across classes through cohorting improves control (SI, subsection 6.9). Despite detecting more cases, regular testing leads to a small increase in student-days lost, <6.6 (6.4-6.8) times the number of days lost with the basic strategy and about 63-80% less than reactive class closure, as isolation is only applied to detected cases. The cost-benefit analysis shows that for all regular testing strategies, the cost expressed by person-days lost remains low, even when the benefit becomes high, for a range of different epidemic conditions (Figure 4b,c). Strategies based on class closures do not manage to reach a high benefit, even at large cost. Reactive screening limits days lost but with a negligible impact on viral circulation. Closing the class at each case detected by regular testing improves case reduction but at the cost of increased absence from school. Findings were robust against changes in detection rates and test sensitivity (SI, subsections 7.4-7.5).

**Figure 4.**
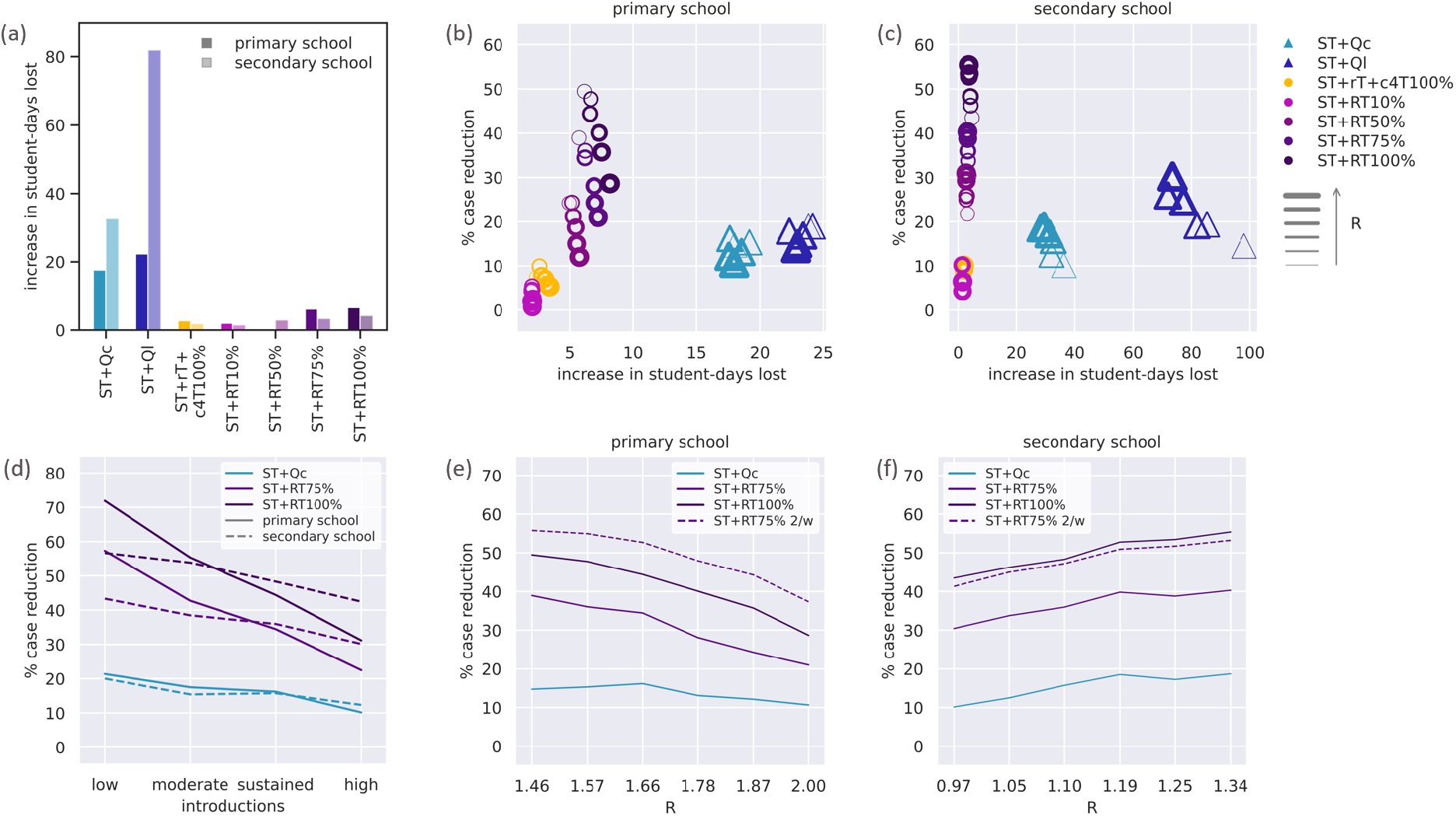
Cost-benefit of regular testing in educational environments and impact of introductions and effective reproductive number. (a) Predicted increase in student-days lost with respect to symptom-based testing (ST) for different protocols in the primary (solid bars) and the secondary (lighter color bars) schools. Intervention protocols are: symptom-based testing with reactive quarantine of the class (ST+Qc); symptom-based testing with reactive quarantine of the class level (ST+Ql); symptom-based testing and reactive screening of the class (+1d from detection), followed by a control screening (+4d) with adherence α=100% (ST+rT+c4T100%); symptom-based testing coupled with regular testing (ST+RTα%) with adherence α 10%, 50%, 75%, 100%. For regular testing, results for weekly frequency are shown. Simulations are parameterized with sustained introductions and the estimated effective reproductive number for the Delta variant, R=1.66 in the primary and R=1.10 in the secondary school, accounting for differences in vaccination coverage. R refers to the ST+Qc protocol with mask mandate, corresponding to the conditions applied in France when data used for the inference were collected. (b) Predicted percentage of reduction in the number of cases vs. predicted increase in student-days lost in the primary school. Both quantities are computed relatively to the basic strategy (symptom-based testing, ST). Each point in the plot corresponds to a protocol (color-coded) and to a value of R (coded with the thickness of the border) in the range 1.46-2.00. Simulations are parameterized with sustained introductions. (c) As panel b, for the secondary school, with R in the range 0.97-1.34. Simulations are parameterized with sustained introductions. (d) Predicted median percentage of reduction in the number of cases achieved by selected protocols with respect to symptom-based testing (ST) as a function of the introductions. Solid lines refer to the primary school, and dashed lines to the secondary school. Regular testing is performed weekly. Simulations are parameterized with the estimated effective reproductive number for the Delta variant (R=1.66 in the primary and R=1.10 in the secondary school). (e) Predicted median percentage of reduction in the number of cases achieved by selected protocols with respect to symptom-based testing (ST) in the primary school as a function of the effective reproductive number R in the range 1.46-2.00. Solid lines refer to weekly RT, dashed line to twice-weekly RT. (f) As in panel e for the secondary school and R in the range 0.97-1.34. In panel e and f simulation are parameterized with sustained introductions.

Higher incidence in the community (increasing the expected introductions at school), and larger reproductive numbers (increasing within-school transmission) reduce the benefit of weekly testing in primary schools, thus requiring increased adherence or frequency (Figure 4d,e). The impact of introductions is milder in the secondary school, due to vaccination (Figure 4f). Moreover, increasing R in this setting would increase the benefit of regular testing, contrary to the primary school case. This is due to a bell-shaped dependence of the infection prevention capacity of regular testing vs. R (SI, subsection 6.10): in low-transmission conditions, only few cases are present even for ST, so that additional protocols yield marginal benefit; as transmission increases from small values (the secondary school case, where R is small thanks to vaccination), efficiency increases; in high-transmission conditions, instead, case prevention is hindered by too many infections generated between successive screenings, and efficiency decreases as transmission increases (the primary school case, with high R because of unvaccinated children). Changes in epidemiological parameters (transmissibility, susceptibility) yield changes in R and consequently in protocols’ efficiencies, but protocols’ ranking according to their benefit remains robust (SI, subsections 7.1-7.3).

Benefits and costs of regular testing remain stable when vaccination coverage of teachers increases from 60% to 100% (Figure 5a and SI, subsections 6.5, 7.6). Increasing vaccination coverage in students, both in primary and secondary schools, is a strong protective factor against school outbreaks (Figure 5b,c,d), expected to reduce the epidemic size by 38% with 20% coverage in children and by 75% with 50% coverage, without intervention (i.e. with ST) and with respect to non-vaccination (Figure 5d, Figure S32). Regular testing would provide an important supplementary control, especially while rolling out vaccination campaigns in primary schools: weekly screening 75% of the non-vaccinated students would additionally reduce cases by 36% (32-39%) with 20% coverage in children, and by 23% (20-26%) with 50% coverage, without impacting class closure (Figure 5e). The minimum vaccination coverage to reduce the benefit of regular testing to 20% case reduction or below increases with R; for R between 1.6 and 2 the required coverage stabilizes around 55-60% (Figure 5f).

**Figure 5.**
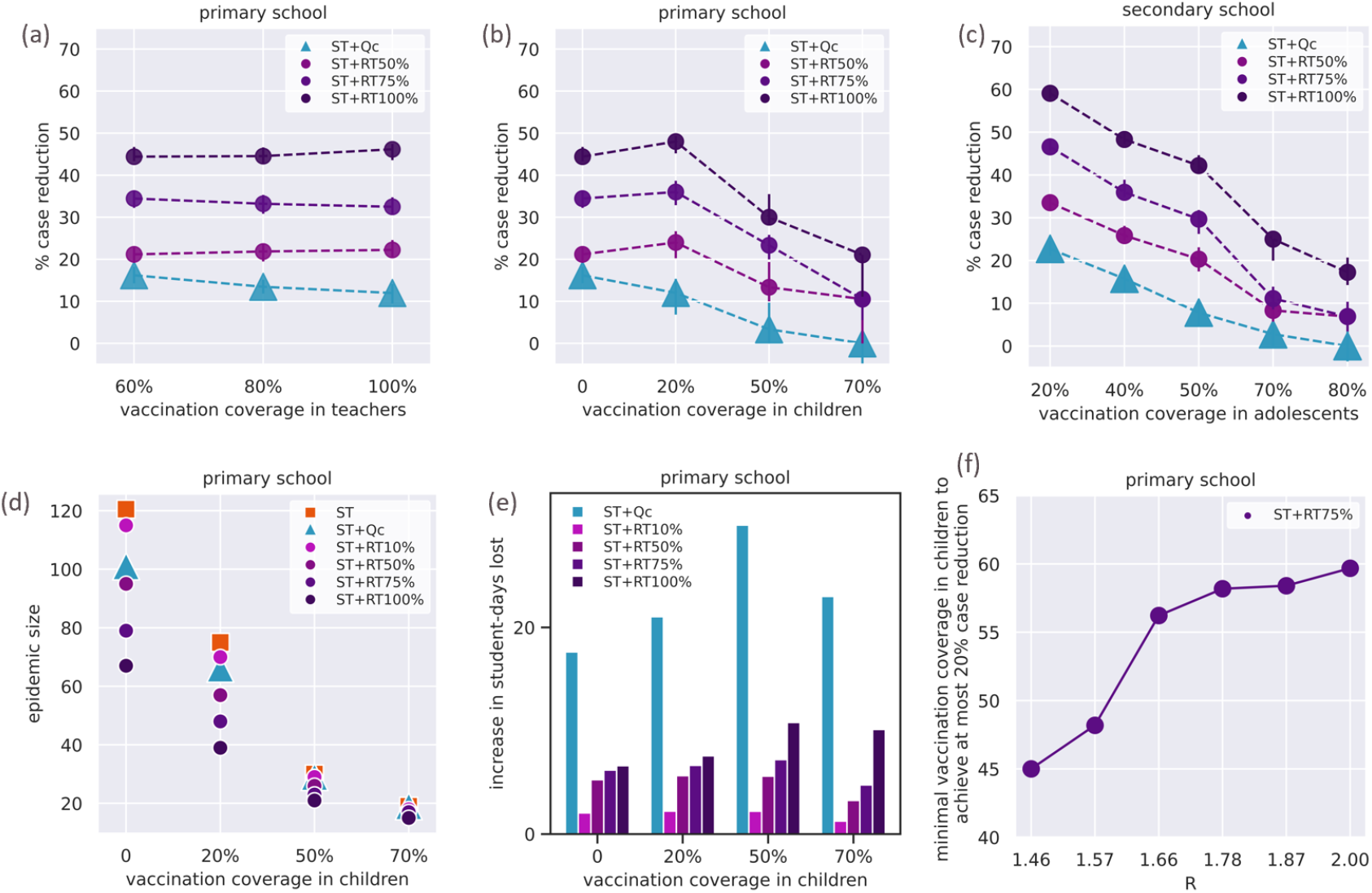
Impact of vaccination coverage. (a) Predicted percentage of reduction in the number of cases achieved by selected protocols as a function of the vaccination coverage in teachers in the primary school. The case reduction is computed relatively to the basic strategy (symptom-based testing, ST). Intervention protocols are: symptom-based testing with reactive quarantine of the class (ST+Qc); symptom-based testing coupled with regular testing (ST+RTα%) with adherence α=50%, 75%, 100%. For regular testing, results for weekly frequency are shown. (b) As in panel a, as a function of vaccination coverage in children. (c) As in panel a, for the secondary school, as a function of vaccination coverage in adolescents. (d) Predicted final epidemic size over the timeframe of 90 days vs. the vaccination coverage in children in the primary school for selected protocols. In addition to the protocols considered in panel a, the panel also shows: symptom-based testing (ST); symptom-based testing coupled with regular testing with 10% adherence (ST+RT10%). (e) Predicted increase in student-days lost for selected protocols as a function of the vaccination coverage in children in the primary school. The increase in days lost is computed relatively to the basic strategy (symptom-based testing, ST). The same selection of protocols is shown as in panel d. (f) Minimal vaccination coverage in children above which weekly testing with 75% adherence (ST+RT75%) in the primary school has at most a benefit of 20% case reduction, as a function of the effective reproductive number R. Simulations are parameterized with sustained introductions. In panels a-e, simulations are parameterized with sustained introductions and the estimated effective reproductive number for the Delta variant (R=1.66 in the primary and R=1.10 in the secondary school). In all panels, R refers to the ST+Qc protocol with mask mandate, corresponding to the conditions applied in France when data used for the inference were collected.

## DISCUSSION

Safely maintaining schools open during the COVID-19 pandemic is a matter of controversial debate and relatively limited knowledge from the field. Using screening data from schools during the 2021 spring wave in France and empirical contact data, our study provides the first estimate of transmissibility in school settings, suggesting that contacts within schools increase SARS-CoV-2 transmission potential compared to the community. As countries in Europe face a new wave due to the Delta variant [13], protocols at school remain a central issue in the midst of vaccine hesitancy and the novel threat of the Omicron variant [14]. Our analysis indicates that regularly screening the school population is efficient in preventing infections while reducing absence from school, especially in settings where the school population is not yet vaccinated or coverage is low to moderate.

We estimated a higher transmissibility in the school compared to the community during the Alpha 2021 spring wave in France. This suggests that repeated contacts in dense classrooms, with mask mandate except during sport and lunch, favor transmission in absence of screening protocols, with potentially high overdispersion [26]. These findings align with available evidence of increased transmission in the population if schools are open [2], [5]. In absence of vaccination, secondary school students are predicted to infect on average a larger number of individuals compared to primary school students, consistent with observations [2], due to age-specific epidemiological properties and contact patterns. However, the more contagious Delta variant and the absence of protection from vaccination currently put children at higher risk. A disproportionately higher viral circulation is observed in children that is further sustained by transmission at school (estimated primary-school-specific R in the range 1.46-2.00 for Delta), resulting in a higher risk of infection for students’ household members [27] and a rapid transmission in the community [28]. Even when vaccination coverage brings school-specific R below 1 (as estimated e.g. in secondary schools in France with 77% vaccinated adolescents; SI, section 4.4), the predicted highly-overdispersed offspring distribution suggests that –together with highly likely extinctions– chains of transmissions in schools are relatively rare but possible.

Using the estimated school-specific transmission rate for Delta and a range of realistic epidemic conditions (introductions, seasonality, vaccination coverage), we found that regular testing with large enough adherence provides an optimal balance in controlling school outbreaks while maintaining schools open. This is consistent with results showing that twice-weekly testing in England helped to control within-school transmission in secondary schools [12]. Adherence is however critical, suggesting that at least ¾ of non-vaccinated individuals should participate to weekly testing to achieve a considerable case reduction. This was not achieved in the pilot screenings in the 2021 spring in France, despite schools mainly participated once. Implementing regular testing in the current context of increasing viral circulation due to the Delta variant [13] should consider improving strategies for the communication and engagement of the school community to considerably boost participation and maintain it over time. This would ensure the school functioning while vaccination campaigns may soon roll out in children.

Our findings corroborate previous numerical evidence on the value of regular testing in preventing infections [9]– [11]. In addition to prior work, our study integrates empirical face-to-face proximity data allowing us to quantify individual-level variation in SARS-CoV transmission. It also provides a cost-benefit analysis comparing multiple protocols and evaluating the key role of adherence in the context of partially vaccinated school populations.

Reactive class closure is highly costly in terms of student-days lost, despite detecting a case is rarer in younger individuals [3]. It also has a limited value in epidemic control, as other classes may be already affected due to unobserved introductions from the community or silent spreading within the school. This second effect becomes particularly important when between-classes mixing is higher, as observed in the primary school. Cohorting that reduces contacts between classes remains therefore an important component of school protocols, in support to screening. While regular testing is able to detect more cases than symptom-based detection, it keeps days lost low for two main reasons. First, isolation is only applied to cases during their infectious period, being therefore more targeted than class quarantine. Second, detecting cases that otherwise go unnoticed helps control the epidemic, breaking the chains of transmission and preventing further diffusion. As a consequence, the overall time spent in isolation is also reduced. Reactive screening, instead, would leave many cases undetected even when retesting a few days after. The iterative nature of the regular testing is key to ensure control over time. Under conditions of high transmissibility or high incidence in the community, complementary efforts should be put in place to counteract the decrease in efficiency of regular testing (e.g., through vaccination, cohorting, and strengthening protocols with higher adherence and frequency of testing), next to the use of masks and ventilation.

Increasing vaccination in teachers protects them from infection and symptomatic disease [21], but yields limited protection for the school population, even under full coverage. This results from the small number of teachers and the observed lower rate of interaction they have with students, and it is confirmed even when community incidence in adults is much higher than in the student age classes. Extending vaccination to students is needed to achieve a collective benefit, reducing the likelihood and size of school outbreaks. In these conditions, regular testing would bring a supplementary control whose application should be evaluated in light of resources, logistics, adherence, and epidemic conditions. Regular testing remains however critical in zero to moderate coverage situations, as it would prevent a substantial portion of undetected infections, with a direct impact to the school environment, reducing the number of infections and long-COVID in children [29], and an indirect impact on the community, protecting students’ contacts [27].

This study has a set of limitations. First, it focuses on two school settings for which empirical contact data were available, but contacts in other schools may be different, depending on the structure of curricula and the organization of activities. Findings on the efficiency of regular testing and vaccination are however robust across a range of epidemic conditions and synthetic contact patterns, and can thus inform on the choice of strategies to safely keep schools open. Second, the study focuses on school outbreaks and it does not assess the impact that these strategies will have on the viral circulation in the community. Third, our protocols’ analysis focuses on the Delta variant. Preliminary data suggests that the Omicron variant has a potential for increased transmissibility compared to Delta [14]. By exploring up to 60% increase in transmissibility of our Delta estimate, our findings reinforce the need for vaccination in the school population to maintain schools open.

In the 2021-2022 winter months, COVID-19 epidemic will likely continue to pose a risk to the safe opening of schools. Regular testing remains a key strategy to epidemic control in school settings with zero to moderate vaccination coverage, all the while minimizing days lost.

## Data Availability

The data that support the findings of this study are openly available at the references cited.

## ACKNOWLEDGMENTS

We thank the Assistance Publique - Hôpitaux de Paris, Santé publique France, Niel Hens, Pieter Libin, Julia Bielicki, Pascal Crepey, and Raphaëlle Métras for useful discussions; Philippe Vanhems, Elisabeth Bothello-Nevers, Olivier Epaulard, Jean Beytout, Annabelle Ravni and the Academie of the Auvergne-Rhône-Alpes region for the school screening initiatives; the Ministry of National Education for supporting the surveillance activities. This study was partially funded by: ANR projects COSCREEN (ANR-21-CO16-0005) and DATAREDUX (ANR-19-CE46-0008-03); ANRS project EMERGEN (ANRS0151); EU H2020 grants MOOD (H2020-874850) and RECOVER (H2020-101003589); EU HORIZON grant VERDI; REACTing COVID-19 grant.

